# Detection Of Genomic Variants Of SARS-CoV-2 Circulating In Wastewater By High-Throughput Sequencing

**DOI:** 10.1101/2021.02.08.21251355

**Authors:** Alba Pérez-Cataluña, Álvaro Chiner-Oms, Enric Cuevas-Ferrando, Azahara Díaz-Reolid, Irene Falcó, Walter Randazzo, Inés Girón-Guzmán, Ana Allende, María A. Bracho, Iñaki Comas, Gloria Sánchez

## Abstract

The use of SARS-CoV-2 metagenomics in wastewater can allow the detection of variants circulating at community level. After comparing with clinical databases, we identified three novel variants in the spike gene, and six new variants in the spike detected for the first time in Spain. We finally support the hypothesis that this approach allows the identification of unknown SARS-CoV-2 variants or detected at only low frequencies in clinical genomes.

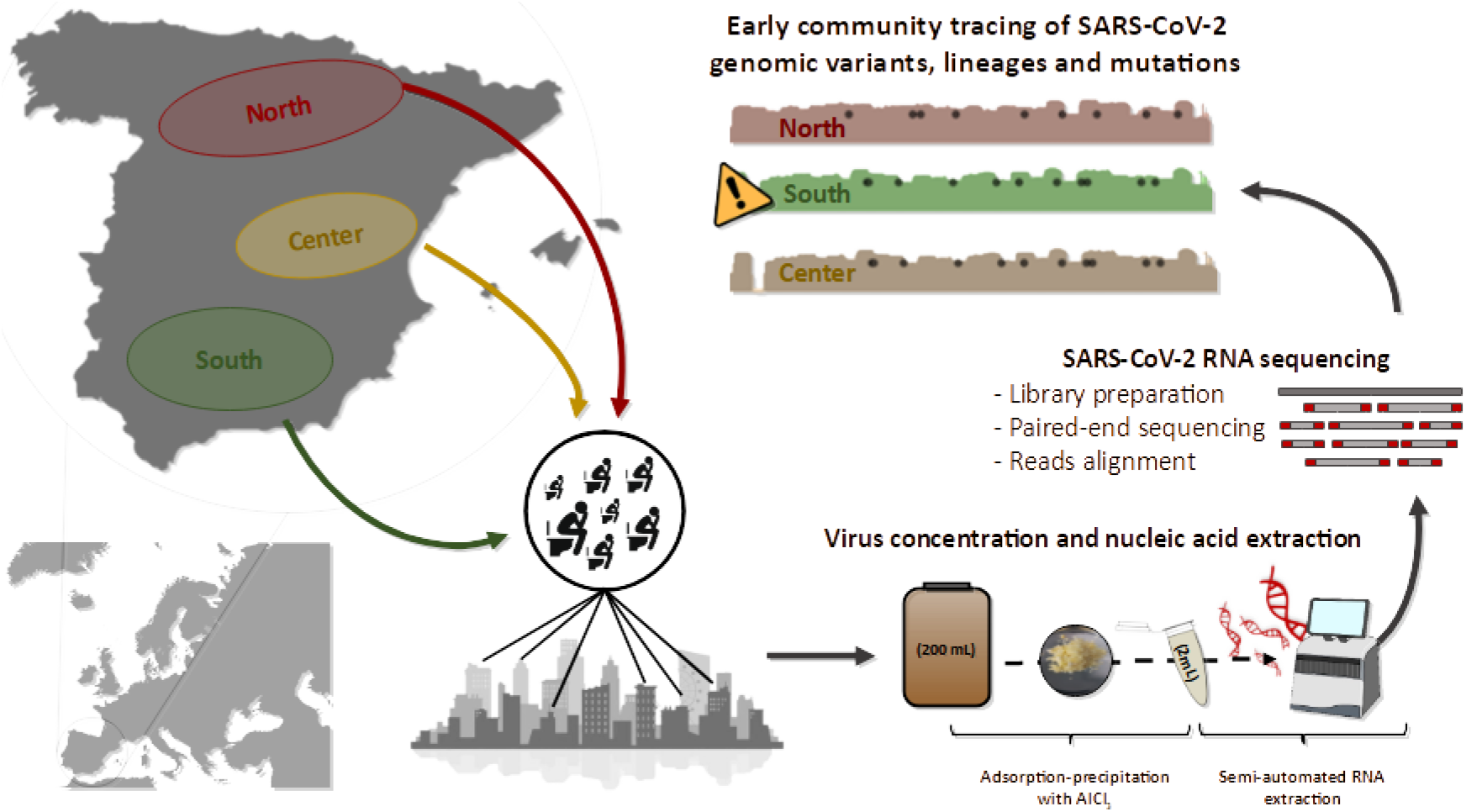

---

The appearance of SARS-CoV-2 and its rapid spread worldwide is causing serious human and economic loss. Transmission of the virus occurs mainly through aerosol and respiratory secretions^1^, but it has also been seen that, due to its replication capacity in the gastrointestinal tract^2^, it is excreted in feces and urine, as previously reported for Severe Acute Respiratory Syndrome (SARS) and Middle-East Respiratory Syndrome (MERS) viruses. For this reason, it has been possible to detect the genetic material of the virus in the feces of both symptomatic and asymptomatic people^3^. These findings have led to the use of wastewater in SARS-CoV-2 monitoring. As with other pathogens, the use of Wastewater-Based Epidemiology (WBE) is a very useful tool for large-scale epidemiological control^4–7^. One of the reasons for the success of WBE is that wastewater samples are a non-invasive and inexpensive source of information to investigate the spread of different genetic variants of SARS-CoV-2 within a community. It provides real-time information on the circulating strains of SARS-CoV-2, which is essential for the development of vaccines and drugs. This is particularly relevant in view of the current situation where the worldwide population is being vaccinated against specific SARS-CoV-2 strains. Massive sequencing techniques allow us to analyze a large number of SARS-CoV-2 genomes, including those present in symptomatic and asymptomatic persons. Through the analysis of sequences, it is possible to detect low-frequency variants (LFV) and to know the gene variants that are circulating at a certain time and place^8,9^. These analyses will make it possible to detect the entry of new lineages into the population, as well as the appearance of polymorphic sites^8,9^.

In this work, 40 grab samples were collected from April to October 2020 from 14 wastewater treatment plants (WWTPs) located in three geographical regions (north: n=5, center: n=21, and south: n=14) within the Spanish peninsula (Table S1). For each sample, 200 mL of influent wastewater samples were concentrated following an aluminum-based adsorption precipitation method^10,11^. Nucleic acid was extracted from wastewater concentrates using an automated method, the Maxwell RSC Pure Food GMO and Authentication Kit (Promega) with slight modifications^12^. SARS-CoV-2 nucleic acid was detected by RT-qPCR using One Step PrimeScript™ RT-PCR Kit (Perfect Real Time) (Takara Bio, USA) targeting three genomic regions of the virus corresponding to N1 region of the nucleocapsid gene (N1), the envelope gene (E), and IP4 region of the RNA-dependent RNA polymerase gene (IP4) using primers, probes and conditions previously described^13–15^. Genomic sequencing of SARS-CoV-2 from wastewater samples was carried out following ARTIC protocol version 3 for retrotranscription and amplification by multiplex PCR (https://www.protocols.io/view/ncov-2019-sequencing-protocol-v3-locost-bh42j8ye). Sequencing libraries were built using the Nextera DNA Flex Library Prep kit (Illumina) and sequenced on the Illumina MiSeq platform by paired-end reads (2×200 bp). Raw reads were cleaned for adaptors and low quality nucleotides by using cutadapt software^16^ and *reformat*.*sh* from bbmap (sourceforge.net/projects/bbmap/), respectively. Nucleotides with Phred score lower than 30 were discarded. Clean reads were aligned to the genome of SARS-CoV-2 isolate Wuhan-Hu-1 (MN908947.3) using the Burrows-Wheeler Aligner v0.7.17-r1188^17^ and indexed by samtools^18^. For the analysis of genomic coverage for each sample, only nucleotides with at least 20X depth were taken into account. Nucleotide variants of SARS-CoV-2 isolate Wuhan-Hu-1 (MN908947.3) were detected with the aligned reads using *mpileup* from samtools^19^ and the command *variants* of ivar software^20^. For the assumption of one variant, at least a 50X depth and quality score higher than 30 were used as cutoff. Alignments were manually curated to avoid nucleotide variants that corresponded to incorrectly trimmed adaptors^8^.

Among the 40 sequenced samples, 28 samples (70%) showed percentages of 20X coverage values higher than 18% of the reference genome length (Figure 1), while the remaining samples ranged from 1.3% to 13.6% (Figure S1). In order to study the potential correlation between viral loads and genome coverage in wastewater samples, Pearson correlation analyses between RT-qPCR outputs (genome copies per liter) versus genome coverage were carried out for each sample. No strong correlations were found for any of the analyzed targets.

**Figure 1.**
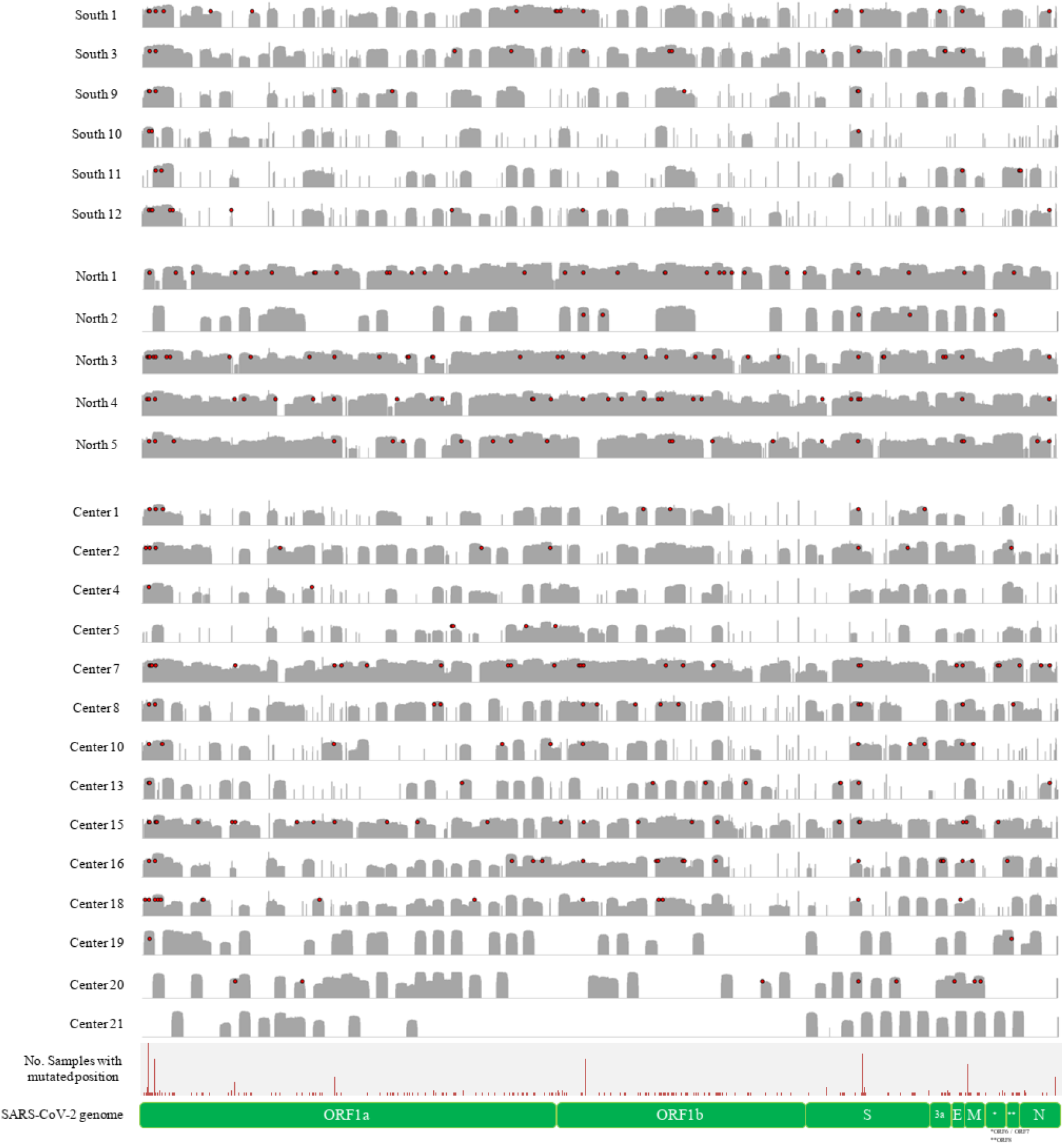
Polymorphic sites (red dots) detected in SARS-CoV-2 genomes from wastewater samples. Genome coverage (>20X) of samples that covered more than 18% of the SARS-CoV-2 isolate Wuhan-Hu-1 genome (MN908947.3) is represented in grey at logarithmic scale (max 4.5 log).

Variant analysis showed a total of 238 nucleotide substitutions and 6 deletions in comparison with the reference genome of SARS-CoV-2 isolate Wuhan-Hu-1 (MN908947.3). Among detected nucleotide variants, 101 polymorphic sites were found in ORF1a polyprotein, 67 in ORF1b, 21 in the spike glycoprotein, 8 in ORF3a, one in the envelope protein, 13 in the membrane glycoprotein, one in ORF7a, 3 in ORF8, 10 in the nucleocapsid gene, one in ORF10, and 12 in intergenic regions (Table 1). In some samples, these nucleotide variants were found in combination with the reference nucleotide present in the genome MN908947.3, referred to as ‘mixed samples’ in Table 1. The percentage of non-synonymous substitutions ranged from 46.1% (in membrane gene) to 100% (in ORF7a and ORF10) (Table 1). Among the nucleotide variants detected in the spike glycoprotein (n=21), 13 of them corresponded to non-synonym substitutions already described (https://mendel.bii.a-star.edu.sg) with the exception of 3 variants that corresponded to the amino acid substitutions G648V, A893T, and L1152S. Seven out of 13 polymorphic sites detected in the spike glycoprotein were not previously described in Spanish genomes (Table 2) according to the database of the Agency of Science, Technology and Research of Singapore (https://mendel.bii.a-star.edu.sg). However, three of these nucleotide changes (amino acid substitutions G404V, G648V, and S884F) have been found at low frequencies among the reads obtained in the sequencing of Spanish genomes from clinical samples. Regarding nucleotide deletions, a total of 6 deletions were found, 4 of them in the ORF1a region (ΔL21/Q22/V23, ΔG82/H83/V84, ΔV84/M85/V86, Δ141K/142S/143), one in the spike glycoprotein (ΔK385), and one in the ORF3a (from amino acid G11 to I20).

**Table 1.**
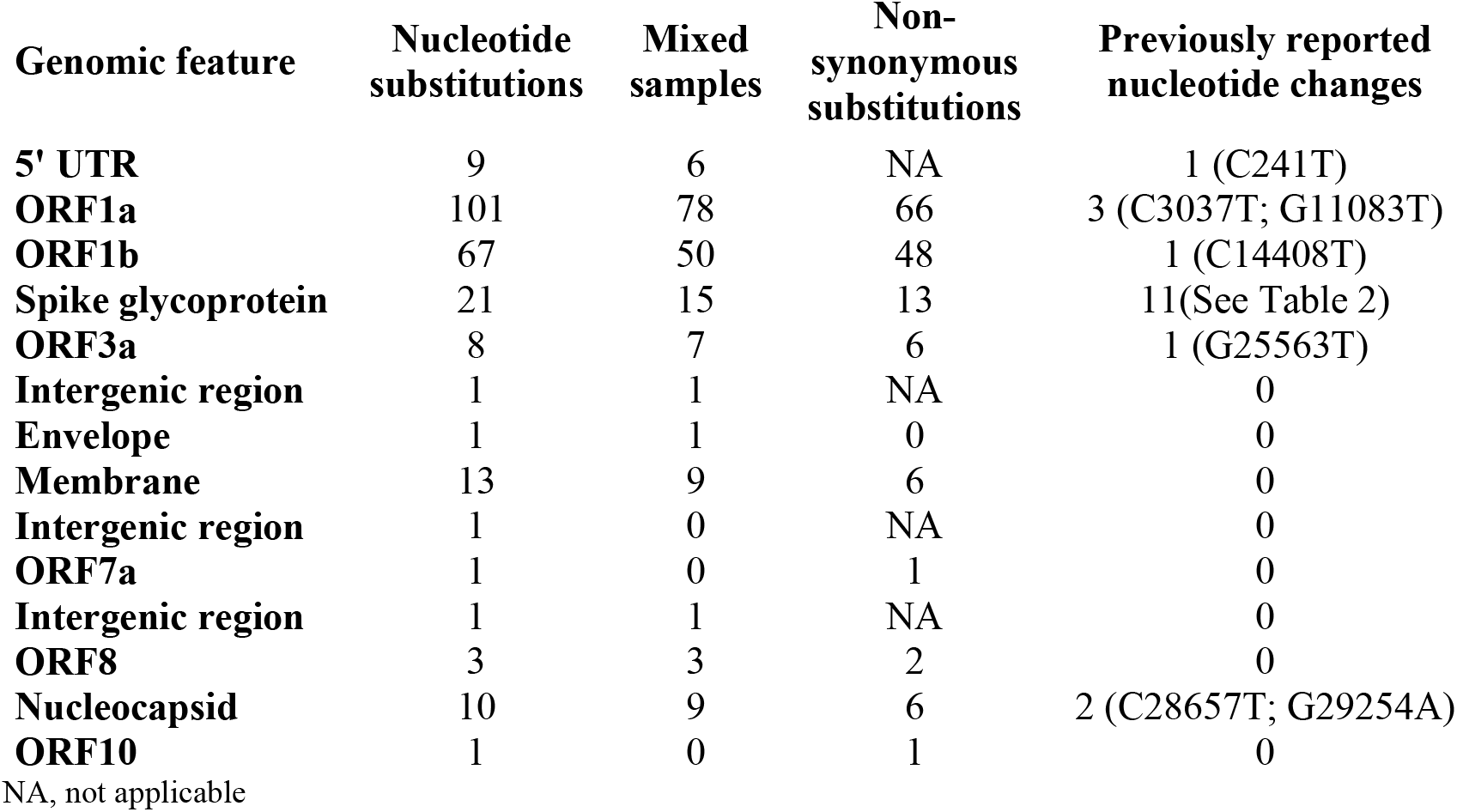
Overview of the nucleotide substitutions detected in SARS-CoV-2 genomes from wastewater samples (n=40) as compared to the SARS-CoV-2 isolate Wuhan-Hu-1 reference genome (MN908947.3). Mixed samples related to the number of samples showing nucleotides according to reference sequence and variant.

**Table 2.** Non-synonymous nucleotide substitutions detected in the spike glycoprotein region as compared to the SARS-CoV-2 isolate Wuhan-Hu-1 reference genome (MN908947.3). Reference and alternative depth relate to the percentage of the total depth that corresponded to the nucleotide present in the reference genome MN908947.3 and the alternative nucleotide, respectively.

| Position in<br>MN908947.3 | No.<br>Samples<br>with<br>reference<br>nt | No.<br>Samples<br>with<br>alternative<br>nt | Nucleotide<br>change | %<br>reference<br>deep | %<br>alternative<br>deep | Amino<br>acid<br>change | Previously<br>described<br>in Spain |
| --- | --- | --- | --- | --- | --- | --- | --- |
| **22227** | 0 | 3 | C22227T | 0 | 100 | A222V | Yes |
| **22676** | 1 | 1 | G22676A | 32 | 68 | A372T | No |
| **22773** | 1 | 1 | G22773T | 83 | 17 | G404V | No* |
| **23162** | 1 | 1 | G23162T | 84 | 16 | V534F | Yes |
| **23403** | 0 | 18 | A23403G | 0 | 100 | D614G | Yes |
| **23477** | 2 | 3 | G23477A | 56-82 | 44-100 | G639S | Yes |
| **23487** | 1 | 1 | T23487G | 41 | 59 | V642G | No |
| **23505** | 1 | 1 | G23505T | 55 | 45 | G648V | NA* |
| **24213** | 1 | 1 | C24213T | 83 | 17 | S884F | No* |
| **24239** | 1 | 1 | G24239A | 84 | 16 | A893T | NA |
| **25017** | 1 | 1 | T25017C | 85 | 15 | L1152S | NA |
| **25047** | 1 | 1 | C25047G | 83 | 17 | P1162R | No |
| **25081** | 0 | 1 | T25081A | 0 | 100 | N1173K | No |

NA, Not applicable, first time described; nt, nucleotide
\*Detected in low frequencies in Spanish clinical genomes.

Nucleotide variants were detected in positions that define SARS-CoV-2 clades, according to the classification of nextstrain.org, even though all the samples carried sequences corresponding to clade 20A (nucleotide substitutions C241T, C3037T, C14408T, and, A23403G). However, samples C1 and C10 showed mixed sequences of clades 20A and 20C at genomic position 25563 (G25563T). Unfortunately, the other nucleotide position that defines clade 20C (position 1059) was not sequenced in sample C10 and presented a low coverage in sample C1, avoiding the further verification of nucleotide substitution (C1059T) at this position.

These results confirm the potential of sewage sequencing to detect clades and new mutations and variants of SARS-CoV-2, which is of utmost relevance for the monitoring efforts for emerging vaccine-escape SARS-CoV-2 mutants in the forthcoming post-vaccination era. Additionally, genomic sequencing of viruses found in wastewater, yields complementary results to those of clinical laboratories, as has been demonstrated with the three novel nucleotide changes in the spike gene identified in wastewater, or the ones initially detected in low number of reads on genomes from clinical specimens and confirmed in wastewater samples. Nevertheless, the different coverage of the genome within individual samples suggests that analysis through massive sequencing focused on genomic regions of interest, such as the spike or clade-defining nucleotide positions, would allow a more robust characterization of the genomic variants spread in a defined geographical area or community.

## Data Availability

Data availability under demand

## Acknowledgements

This study was supported by projects “VIRIDIANA” (AGL2017-82909/ AEI/FEDER, UE) funded by Spanish Ministry of Science, Innovation and Universities; CSIC (202070E101), and Generalitat Valenciana (Covid_19-SCI). EC-F is recipient of a predoctoral contract from the MICINN, Call 2018. We acknowledge NILSA, FACSA, and MITECO for authorizing the sampling. The authors thank Agustin Garrido Fernández, and Andrea Lopez de Mota for their technical support.

## Contributions

AP-C, AC-O, EC-F, AD-R: Investigation, formal analysis, writing, and reviewing. IF-F, IG-G: Resources, writing, and reviewing. WR, AA, MAB, IC: Writing, and reviewing. AP-C, GS: Conceptualization. GS: Funding acquisition, writing, and reviewing. All authors have read and agreed to the published version of the manuscript.

## SUPPLEMENTARY MATERIAL

**Supplementary table S1.** Summary of the analyzed samples and results obtained for detection of SARS-CoV-2 by RT-qPCR, genome coverage (higher than 20X), mean depth of the sample sequencing, and number of nucleotide variants detected. ND, not determined; N1, region N1 of the nucleocapsid gene; E, envelope gene; IP4, region of the RNA-dependent RNA polymerase gene.

| Sample | Sampling date<br>(mm/dd/yyyy) | Ct value<br>(N1)*<sup>13</sup> | Ct value<br>(E)*<sup>14</sup> | Ct value<br>(IP4)*<sup>15</sup> | 20x<br>genome<br>coverage<br>(%) | Mean<br>depth (nt) | No.<br>Variants |
| --- | --- | --- | --- | --- | --- | --- | --- |
| C1 | 8/18/2020 | 29.03 | 31.77 | 34.77 | 43.83 | 297.80 | 7 |
| C2 | 9/8/2020 | 30.40 | ND | ND | 66.61 | 766.60 | 9 |
| C3 | 9/15/2020 | 31.35 | ND | ND | 3.03 | 71.17 | 0 |
| C4 | 9/15/2020 | 31.00 | ND | ND | 43.90 | 292.72 | 2 |
| C5 | 9/22/2020 | 30.97 | ND | ND | 39.15 | 176.42 | 4 |
| C6 | 9/22/2020 | 31.47 | ND | ND | 18.26 | 293.42 | 5 |
| C7 | 9/2/2020 | 31.70 | 30.50 | 32.27 | 94.35 | 1,255.40 | 29 |
| C8 | 9/10/2020 | 32.98 | - | 34.63 | 56.15 | 636.50 | 15 |
| C9 | 9/10/2020 | 31.29 | - | 35.57 | 1.31 | 70.75 | 0 |
| C10 | 10/1/2020 | 32.06 | ND | 35.73 | 42.98 | 625.96 | 11 |
| C11 | 10/1/2020 | 30.98 | ND | 40 | 3.03 | 123.22 | 0 |
| C12 | 10/8/2020 | 30.12 | ND | 34.79 | 8.64 | 132.48 | 0 |
| C13 | 8/26/2020 | 31.80 | 32.44 | 34.03 | 36.89 | 638.74 | 10 |
| C14 | 9/2/2020 | 31.99 | 30.68 | 33.03 | 1.72 | 98.29 | 0 |
| C15 | 9/9/2020 | 31.29 | 32.93 | 32.64 | 82.98 | 759.03 | 25 |
| C16 | 9/29/2020 | 26.59 | ND | 32.89 | 59.45 | 523.76 | 18 |
| C17 | 10/6/2020 | 32.44 | ND | 36.80 | 5.66 | 145.20 | 0 |
| C18 | 10/13/2020 | 33.36 | ND | 34.48 | 60.64 | 363.06 | 14 |
| C19 | 4/6/2020 | 34.53 | ND | ND | 38.37 | 1,942.57 | 2 |
| C20 | 4/6/2020 | 34.75 | ND | ND | 44.68 | 1,882.54 | 8 |
| C21 | 4/6/2020 | 34.67 | ND | ND | 26.33 | 1,671.01 | 0 |
| N1 | 4/30/2020 | 32.43 | ND | ND | 93.06 | 1,566.53 | 33 |
| N2 | 5/5/2020 | 33.08 | ND | ND | 42.99 | 2,003.25 | 5 |
| N3 | 9/3/2020 | 30.79 | 29.96 | 31.80 | 94.80 | 1,663.94 | 34 |
| N4 | 9/10/2020 | 30.40 | 31.61 | 32.17 | 96.81 | 1,401.06 | 28 |
| N5 | 9/17/2020 | 30.72 | ND | 32.76 | 88.77 | 1,504.50 | 20 |
| S1 | 9/1/2020 | 32.57 | 29.36 | 33.51 | 66.38 | 712.84 | 16 |
| S2 | 9/8/2020 | 32.98 | - | 37.33 | 8.45 | 140.17 | 1 |
| S3 | 9/15/2020 | 32.58 | - | 34.19 | 73.60 | 748.27 | 14 |
| S4 | 9/10/2020 | 33.25 | 34.92 | 37.10 | 13.59 | 345.55 | 0 |
| S5 | 9/4/2020 | 30.73 | 31.06 | 34.38 | 21.99 | 266.82 | 3 |
| S6 | 9/11/2020 | 30.96 | - | 35.20 | 11.77 | 69.15 | 0 |
| S7 | 10/2/2020 | 30.44 | - | 37.48 | 1.85 | 110.95 | 0 |
| S8 | 10/7/2020 | 30.13 | - | 31.39 | 1.90 | 82.47 | 0 |
| **S9** | 9/2/2020 | 31.63 | 30.44 | 34.61 | 43.00 | 449.91 | 8 |
| **S10** | 9/9/2020 | 32.67 | - | 34.97 | 24.77 | 356.15 | 3 |
| **S11** | 9/8/2020 | 31.78 | - | 34.67 | 32.45 | 467.41 | 5 |
| **S12** | 9/15/2020 | 31.58 | - | 33.94 | 43.48 | 549.82 | 12 |
| **S13** | 9/29/2020 | 31.49 | - | 36.02 | 18.09 | 233.10 | 5 |
| **S14** | 10/14/2020 | 31.17 | - | 33.77 | 10.25 | 254.33 | 0 |

\*Average values of two replicates
<sup>13</sup>CDC, 2020.
<sup>14</sup>Corman et al., 2020
<sup>15</sup>Institut Pasteur, Paris.

**Supplementary Figure S1.**
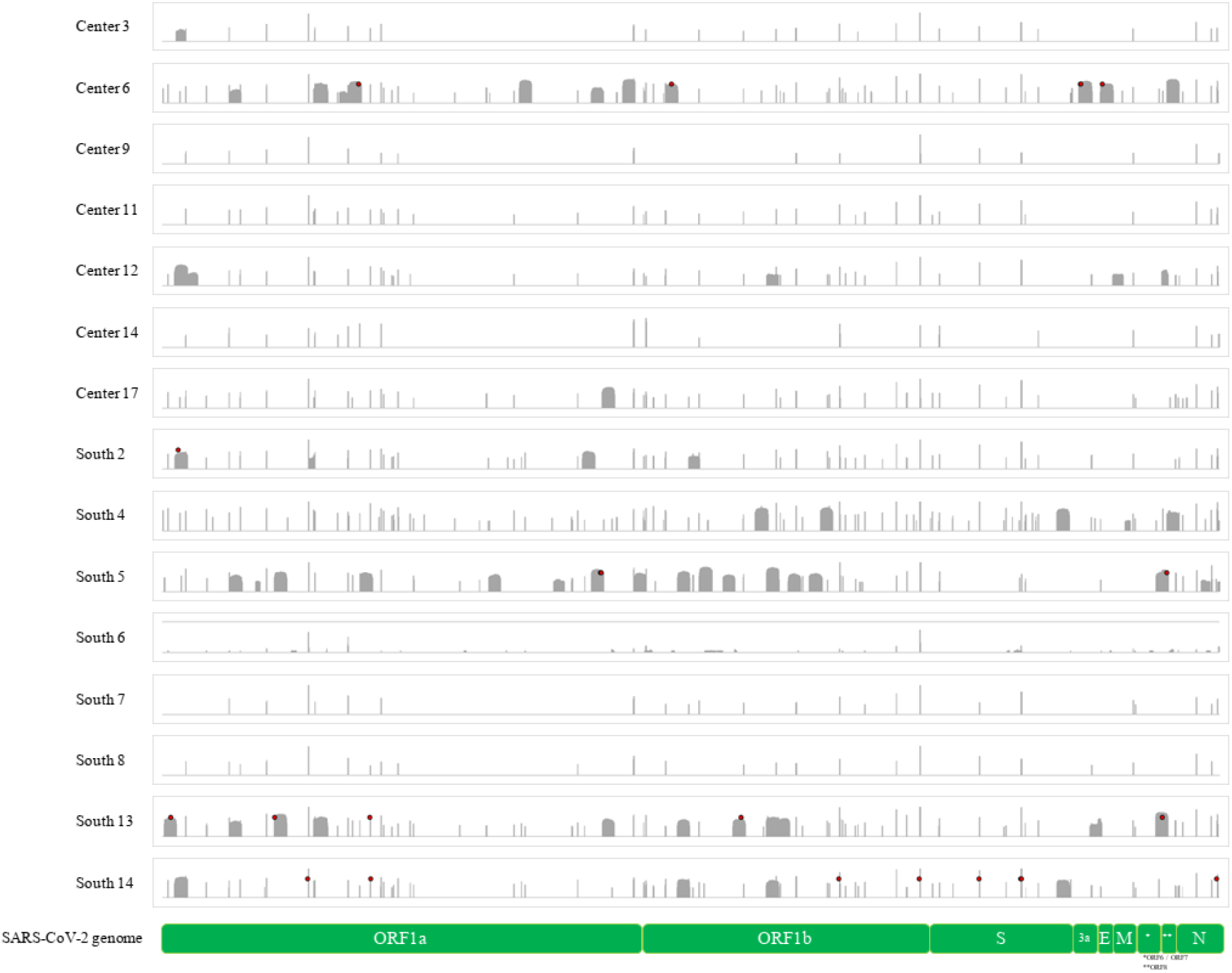
Representation of the genome coverage (>20X) in logarithmic scale (max 4.5 log) reached by samples that covered less than 18% of the SARS-CoV-2 isolate Wuhan-Hu-1 genome (MN908947.3).

## References

1. Chan, J. F. W. et al. Genomic characterization of the 2019 novel human-pathogenic coronavirus isolated from a patient with atypical pneumonia after visiting Wuhan. Emerg. Microbes Infect. 9, 221–236 (2020).

2. Xiao, F. et al. Evidence for Gastrointestinal Infection of SARS-CoV-2. Gastroenterology 158, 1831-1833.e3 (2020).

3. Polo, D. D. et al. Making waves: Wastewater-based epidemiology for SARS-CoV-2 – Developing robust approaches for surveillance and prediction is harder than it looks. Water Res. 186, 116404 (2020).

4. Heijnen, L. & Medema, G. Surveillance of influenza A and the pandemic influenza A (H1N1) 2009 in sewage and surface water in the Netherlands. J. Water Health 9, 434–442 (2011).

5. Hellmér, M. et al. Detection of pathogenic viruses in sewage provided early warnings of hepatitis A virus and norovirus outbreaks. Appl. Environ. Microbiol. 80, 6771–6781 (2014).

6. Santiso-Bellón, C. et al. Epidemiological Surveillance of Norovirus and Rotavirus in Sewage (2016-2017) in Valencia (Spain). Microorganisms 8, (2020).

7. Cuevas-Ferrando, E., Randazzo, W., Pérez-Cataluña, A. & Sánchez, G. HEV Occurrence in Waste and Drinking Water Treatment Plants. Front. Microbiol. 10, 2937 (2019).

8. Nemudryi, A. et al. Temporal Detection and Phylogenetic Assessment of SARS-CoV-2 in Municipal Wastewater. Cell Reports Med. 1, 100098 (2020).

9. Crits-Christoph, A. et al. Genome Sequencing of Sewage Detects Regionally Prevalent SARS-CoV-2 Variants. MBio 12, (2021).

10. AAVV. “Section 9510 D. Virus concentration by aluminum hydroxide adsorption-precipitation, chapter detection of enteric viruses”. in Standard Methods for the Examination of Water and Wastewater (ed. E. W. Rice, R. B Baird, and A. D. E.) (Denver, CO: American Water Works Association, 2018).

11. Randazzo, W. et al. SARS-CoV-2 RNA in wastewater anticipated COVID-19 occurrence in a low prevalence area. Water Res. 181, 115942 (2020).

12. Pérez-Cataluña, A. et al. Comparing analytical methods to detect SARS-CoV-2 in wastewater. Sci. Total Environ. 758, 143870 (2021).

13. CDC. CDC 2019-novel coronavirus (2019-nCoV) real-time RT-PCR diagnostic panel. https://www.fda.gov/media/134922/download. (Accessed Oct. 2020).

14. Corman, V. M. et al. Detection of 2019 novel coronavirus (2019-nCoV) by real-time RT-PCR. Eurosurveillance 25, 2000045 (2020).

15. Institut Pasteur, P. Protocol: Real-Time RT-PCR Assays for the Detection of SARS-CoV-2. Available online: https://www.who.int/docs/default-source/coronaviruse/real-time-rt-pcr-assays-for-the-detection-of-sars-cov-2-institut-pasteur-paris.pdf?sfvrsn=3662fcb6_2 (Accessed Oct. 2020)

16. Martin, M. Cutadapt removes adapter sequences from high-throughput sequencing reads. nEMBnet.journal 17, 10 (2011).

17. Li, H. & Durbin, R. Fast and accurate short read alignment with Burrows-Wheeler transform. Bioinformatics 25, 1754–1760 (2009).

18. Li, H. et al. The Sequence Alignment/Map format and SAMtools. Bioinformatics 25, 2078–2079 (2009).

19. Li, H. A statistical framework for SNP calling, mutation discovery, association mapping and population genetical parameter estimation from sequencing data. Bioinformatics 27, 2987–2993 (2011).

20. Grubaugh, N. D. et al. An amplicon-based sequencing framework for accurately measuring intrahost virus diversity using PrimalSeq and iVar. Genome Biol. 20, 8 (2019).

